# Triaging and Referring In Adjacent General and Emergency Departments: a six-year follow-up study after a cluster randomised trial

**DOI:** 10.64898/2026.03.21.26348955

**Authors:** Stefan Morreel, Mark Timmermans, Koenraad G Monsieurs, Anthony Pairon, Veronique Verhoeven

## Abstract

**Objectives:** Emergency department (ED) overcrowding is a persistent issue in European healthcare systems. A previous randomized controlled trial (RCT) concerning out of hours care in Antwerp (2019) demonstrated that a nurse-led triage tool, extending the Manchester Triage System (eMTS), could safely redirect low-acuity ED patients to a co-located General Practitioner Cooperative (GPC). This study reports a six-year follow-up assessing long-term efficiency, safety, and sustainability of this intervention.

**Methods:** We performed a retrospective observational analysis of routine clinical data. Patients triaged at the ED and referred to the GPC were identified through electronic health records. Efficiency outcomes included the proportion of ED patients managed at the GPC, the proportion of GPC patients originating from the ED and their clinical characteristics. To assess safety, we analysed rates and characteristics of patients referred back from the GPC to the ED. A detailed case review was conducted for all back-referred patients.

**Results:** Of the 110,941 triaged patients, 6,722 (6.1%) were managed at the GPC, accounting for 11% of all GPC consultations. Diverted patients typically presented with digestive, respiratory, and musculoskeletal complaints and had a clinical urgency which was mostly comparable to the overall GPC population. Only 3% of the patients diverted to the GPC were referred back to the ED, versus 5% of other GPC patients. Most back-referrals (83%) were managed on an outpatient basis; four major and 18 minor triage issues were identified, without evidence of increased morbidity.

**Conclusions:** Six years post-trial, the nurse-led eMTS triage tool remains integrated into routine practice, with increasing efficiency and remaining safety without dedicated research resources nor a post implementation plan. Sustained adoption highlights its clinical feasibility and long-term safety. Future trials on triage and primary care should embed explicit post-trial implementation strategies to promote continuity and scalability of successful healthcare interventions.

**ClinicalTrials.gov Identifier:** NCT03793972

## Introduction

When patients experience acute health problems outside regular office hours, they rely on the out-of-hours (OOH) care system. In Europe OOH primary care is increasingly organised through large-scale General Practitioner Cooperatives (GPCs), whereas hospital emergency departments (EDs) operate in parallel to these GPCs. In Belgium, patients do not need a referral to visit the ED; their choice between ED and primary care is shaped by accessibility, prior experiences, perceived severity of illness, and the patient–GP relationship.[1] A substantial proportion of ED presentations could be managed within primary care[2], however expansion of GPC networks has not led to a decline in ED use.[3]

Overcrowded EDs seek for an efficient collaboration with primary care. A range of physical triage interventions have been suggested to redirect suitable patients from ED to primary care and thus optimise resource use, improve patient flow and reduce clinician workload at the ED.[4] Evidence on efficiency and especially safety in real life is limited.[5]

In this context, a cluster randomised trial in an ED and adjacent GPC in Antwerp[6], conducted from March to December 2019, evaluated whether a triage protocol, adapted from the Manchester Triage System (MTS) could safely redirect a proportion of ED patients to a co-located GPC (the TRIAGE trial). All patients with a national insurance number triaged by a nurse at the ED were included. In this “extended MTS” (eMTS), existing MTS flowcharts were adapted to include the possibility of GPC referral in certain predefined circumstances, as agreed by ED and GPC physicians in a consensus procedure. The eMTS was then integrated into the ED’s computer decision support system. ED nurses in the project followed a twelve-hour training on using the eMTS and communicating the triage outcome to patients. In the actual trial, 13% out of the 6374 included patients in the intervention group were assigned to the GPC; 71% of them (599 patients or 9.5%) were seen at the GPC and four percent were referred back to the ED. Seven percent of the patients seen at the GPC in the study period were referred from the ED.

A process evaluation showed that the intervention was perceived as positive by physicians and nurses, leading to a perceived lower workload at the ED despite the extra efforts of the triage process.[7]

While the immediate impact on patient flow and workload of that and similar interventions is well-documented, evidence regarding long-term sustainability of such initiatives remains limited. In this study we report on the adoption of our study protocol in routine practice and the long-term referral activities at our study site, up to six years after conclusion of the RCT in December 2019.

Efficiency as well as safety are assessed by studying routine clinical data and by performing a case review of patients triaged to the GPC but referred back to the ED after GPC consultation.

## Materials and methods

### Original Study design

The TRIAGE trial was a single centre randomised controlled trial conducted from 01/03/2019 until 31/12/2019. Weekends served as units of randomisation and patients as units of analysis. Patients presenting themselves at the GPC and those arriving at the ED by an ambulance staffed with a doctor or a nurse were excluded.

The MTS (version 3.6) is a validated tool for prioritisation patients in the ED, it is used by nurses worldwide. MTS triage results in a reason for encounter (e.g., abdominal pain in children) and an urgency category ranging from one (immediate care necessary) to five (non-urgent). For the TRIAGE trial, an extended version of the MTS (eMTS) was created. In general, patients in urgency categories four and five were allowed to be assigned to the GPC if they were not in need of hospital care (including lab testing, imaging, suturing and specialist consultation). The nurses were allowed to overrule the result of this automated eMTS assignment using their gut feelings.[16]

During control weekends the assignment was not communicated to the patients, they all remained at the ED. During intervention weekends patients were encouraged to comply with the assignment but were allowed to refuse it.

The intervention was continued after the end of the trial without further control weekends. During the trial, the GPC did not have appointments, patients were handled in order of their presentation. During the COVID-19 pandemic, appointments were introduced and remained afterwards, although the appointment system changed frequently.

### Data collection

#### Routine clinical data study

For the routine clinical data study, no linked records were available as the database used for the original study (containing linked GPC and ED patient data) was no longer updated. GPC data were collected using the electronic health record of the GPC (Mediris Wachtpost 2.55, Mediportal, Belgium) on 09/04/2025. Data were extracted by the GPC manager from the start of the TRIAGE trial (1/3/2019) until 9/2/2025, nearly six years in total. The patient ID was pseudonymised using the SHA-256 algorithm, the database did not contain any other identifiable information. This database contains the routine clinical data for all GPC contacts: demographic data (age, gender, ZIP-code), reason for encounter (International Catalogue of Primary Care (ICPC-2) coded), diagnosis (ICPC-2 coded), GP’s assessment of urgency after consultation (five levels, from “Highly Urgent” to “Not In Need Of OOH Care”) and patient’s origin (walk-in, after telephone contact, after ED triage, leaving ED without triage). Waiting time (in minutes) was calculated as the timestamp of opening the medical record by the GP minus the timestamp of administrative registration at the ED with the exclusion of outliers above 240 minutes (most likely due to late registration not late consultation).

When registering patients, the GPC’s receptionist mandatorily registers the origin of the patient (walk in, after ED triage or after ED but on their own initiative). During the TRIAGE trial, the GPC was only open during weekends, bridging days and holidays. As a response to the COVID-19 pandemic, the opening hours were expanded to weeknights (19.00-8.00); this change was later retained. After 01/01/2023, the GPC was no longer willing to help triaged patients between 00.00 and 6.00.

ED data were extracted using E.care (Mesalvo Turnhout, Belgium) on 08/01/2026. Because of strict privacy regulations, no detailed ED data were available, the only extracted variable was the admission time stamp of all patients registered at the ED. The total number of triaged patients equalled the total number of ED visits during the opening hours of the GPC as all patients were triaged without exceptions.

#### Case review study

For the case review study, the coordinator of the GPC securely transferred an identifiable list of all patients in the referred back group on 25/02/2025 to author MT, head of the study ED. Author MT carefully reviewed all files from patients in this list in the ED’s electronic health records. The following data were extracted (anonymous)119-: demographics (gender, age, ZIP-code), reason for encounter (MTS flowchart), drug prescription (yes/no and specification), drug administration route (oral, intravenous or rectal), hospitalisation (yes/no), hospital procedures (yes/no, defined as procedures generally not used in primary care, such as endoscopy), X-ray (yes/no and specification), other radiology (yes/no and specification), lab test (yes/no and specification), ECG (yes/no), diagnosis (ICPC-2 coded by the authors), retrospective judgement on triage (correct, minor issue or major issue). Minor issues were defined as situations where it might have been better not to triage the patient to the ED, but the patient was not a risk of additional morbidity or mortality (e.g. a delay in treatment for a fractured ankle). Major issues were defined as situations where the loss of time during transfer to the GPC and back to the ED might have led to additional morbidity or mortality (e.g. a delay in treatment of a stroke patient). During the TRIAGE trial, abdominal pain turned out as the eMTS flowchart with the highest risk of referral back to the ED, often with a request to exclude appendicitis. That is why the additional variables Referred Back to Exclude Appendicitis (yes/no) and Appendicitis Surgery (yes/no) were examined as well. In some records information concerning care at other EDs was found, these data were registered as well. All personal data were removed from this list before transfer to the other researchers.

### Studied populations

The *overall GPC population* consisted of all consultations at the GPC during the study period, excluding telephone contacts and home visits. Consultations on bridging days were excluded because during these busy days, the ED was not allowed to divert patients to the GPC.

The *primary outcome group* consisted of all included patients handled at the GPC after ED triage as recorded by the GPC’s receptionist. Patients allocated to the GPC but who never arrived there (either because they insisted on staying at the ED or because they chose to wait or went elsewhere) were not analysed in this group because this group was not identifiable.

The *referred back group* is a subset of the primary outcome: patients allocated to the GPC after triage, handled by the GPC but referred back to the ED by the GP.

### Outcomes

The primary outcomes of this follow-up study were the proportion of triaged ED patients managed at the GPC and the proportion of GPC patients originating from the ED. The former is a more relevant outcome for efficiency measurement but only for the latter (the primary outcome group) individual patient data were available. The nominator for both outcomes is very similar but not exactly the same (as the registrations of ED and GPC contained some errors), the denominator is different.

Secondary outcomes regarding efficiency: characteristics of the primary outcome population as compared to the overall GPC population.

Secondary outcomes regarding safety: proportion of primary outcome patients referred back to the ED after GP consultation and the characteristics of this referred back population.

### Ethics

Ethical clearance waiving individual informed consent was obtained from the ethics committee of Antwerp University Hospital (reference 18/37/410) with an amendment for the current study (reference 3374).

### Analysis

JMP pro version 17 (JMP Statistical Discovery LLC) was used for all analyses. Pearson Chi Square tests, Fisher Exact and unpaired T-tests were used to compare the different study populations.

## Results

### Routine clinical data study

#### Primary outcomes

The proportion of triaged ED patients managed at the GPC was 6.1% (6722/110941) which corresponds to 11% of the GPC population (see Table 1). This proportion decreased during the COVID-19 pandemic and increased afterwards.

**Table 1:**
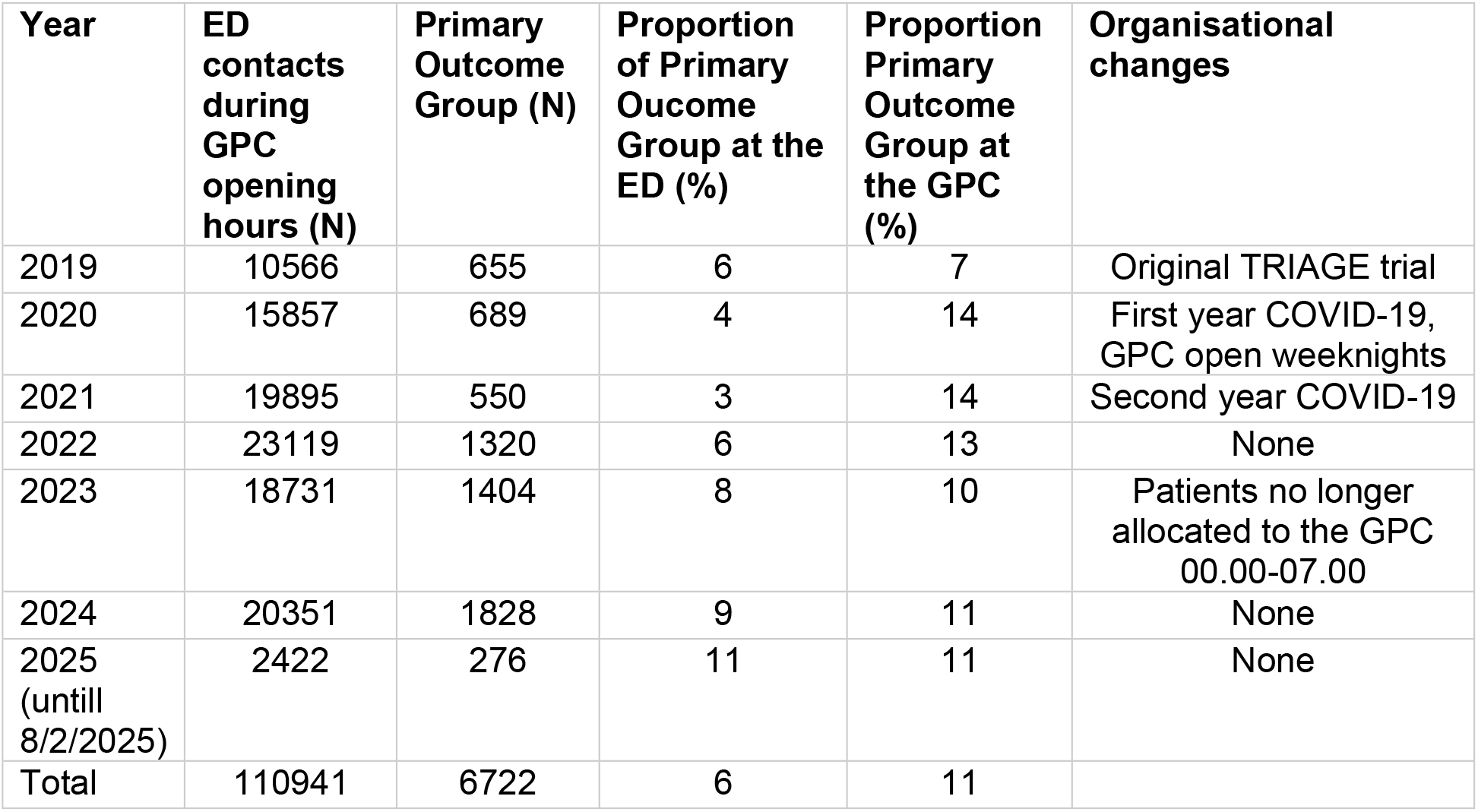
Evolution of the primary outcome over time. ED: Emergency Department, GPC: General Practice Cooperative.

According to the assessment by the GPs, 49% of the primary outcome patients in 2024-2025 did not need OOH medical care (see Table 1). Although this proportion seems high, it is almost equal to the assessment of the overall GPC population (48%). The necessity of urgent care was lower among the primary outcome population; none of them needed highly urgent care (see Table 2).

**Table 2:** GP’s assessment of urgency among the overall GPC population and the primary outcome population. (2024-2025 only).

| GP urgency category | Overall GPC population | % | Primary Outcome Population | % |
| --- | --- | --- | --- | --- |
| Life threatening | 1 | 0% | 0 | 0% |
| Highly Urgent | 8 | 0% | 0 | 0% |
| Urgent | 276 | 2% | 21 | 1% |
| Needs OOH care | 8876 | 51 % | 1043 | 50% |
| Not in need of OOH Care | 8412 | 48 % | 1038 | 49% |

GP: General practitioner, GPC: General Practice Cooperative, OOH = Out of Hours

Patients in the primary outcome group mainly had typical primary care complaints: reasons for encounter and diagnosis mainly concerned the digestive, musculoskeletal and respiratory system (see S1 and S2 tables).

The primary outcome group differed slightly from the overall GPC population in terms of age (means 30 years and 31 years, unpaired T-test P-value <0.01), waiting time (23.5 minutes and 24.4 minutes, T-test p-value <0.01) and gender (53% and 49% female, Chi-square P-value <0.01); they lived outside of the working area of the GPC more often (22% versus 10%, Chi square P-value<0.01).

The mean waiting time of the primary care group was 11 minutes with 98% seen within 45 minutes. Some patients might have had a longer waiting time as the time between triage at the ED and registration at the GPC could not be measured.

### Referred back group

The proportion of the primary outcome patients referred back to the ED was 3% (185/6722) which is lower than among the GPC patients not originating from the ED (5%, 2807/56070, Chi square P-value<0.01). Twelve patients were referred to other EDs nearby (including five children referred to a paediatric hospital), 173 were referred back to the adjacent ED.

Patients triaged to the GPC with digestive, general urinary, and female genital diagnoses were referred back above average whereas skin and ear diagnosis were referred back below average (see Table 3).

**Table 3:** comparison of the proportion of patients referred back to the ED among the primary outcome populations between diagnosis (ICPC-2 chapters).

| Diagnosis ICPC-2 Chapter | Handled at the GPC<br>Referred back to the ED |  |  |  | P-value |
| --- | --- | --- | --- | --- | --- |
|  | N | Row % | N | Row % |  |
| **Digestive** | **1323** | **95%** | **68** | **5%** | **<0.01** |
| Musculoskeletal | 1124 | 98% | 24 | 2% | 0.13 |
| **Skin** | **972** | **99%** | **10** | **1%** | **<0.01** |
| Respiratory | 925 | 99% | 14 | 1% | 0.01 |
| **General** | **761** | **96%** | **35** | **4%** | **<0.01** |
| **Ear** | **536** | **99%** | **3** | **1%** | **<0.01** |
| Neurological | 179 | 96% | 7 | 4% | 0.36 |
| Eye | 177 | 97% | 6 | 3% | 0.64 |
| **Urinary** | **153** | **94%** | **9** | **6%** | **0.05** |
| Psychological | 117 | 100% | 0 | 0% | 0.08 |
| Cardiovascular | 105 | 98% | 2 | 2% | 1 |
| Male genital | 67 | 99% | 1 | 1% | 1 |
| **Female Genital** | **26** | **90%** | **3** | **10%** | **0.04** |
| Blood | 22 | 92% | 2 | 8% | 0.14 |
| Endocrine/Metabolic/Nutritional | 23 | 100% | 0 | 0% | 1 |
| Pregnancy | 11 | 92% | 1 | 8% | 0.28 |
| Social Problems | 12 | 100% | 0 | 0% | 1 |
| All | 6533 | 97% | 185 | 3% |  |

Diagnosis with a p-value <0.05 are shown in bold. P-value for Pearson Chi<sup>3</sup> or when applicable Fisher Exact test. ED= Emergency Department, GPC = General Practice Cooperative, ICPC= International Catalogue of Primary Care)

As compared to the rest of the primary outcome group, the referred back group was similar in age (both 31 years on average), sex (female 49% vs 45%, Chi square P-value 0.33) and living in the working area of the GPC (both 22%).

The referred back group had a mean waiting time at the GPC of 13 minutes with 98% seen within an hour at the GPC.

### Case review study

For the case review study, 185 files were reviewed. For 7 patients (4%), no triage data were found. Most likely these patients were not registered correctly by the GPC’s receptionist and were in fact walk-in patients. These have been excluded from further analysis in this case review study. For 23 patients, only a triage contact was found in the ED files but no ED physician contact nor GPC consultation. These patients either went home without further help or went to another ED (at least 12 see above), another GPC or their own GP later. As we don’t have further data on these patients; they have been excluded from further analyses as well. The remaining 155 were further analysed as the referred back patients (RBPs).

The most common eMTS flow charts for RBPs were: Abdominal Pain in Adults (N=41), Unwell Adult (N=23), Limb Problem (N=14), Unwell Child (N=12), Abdominal Pain in children (N=11), all other flow charts were used less than ten times.

Table 4 provides an overview of the delivered hospital care for RBPs. Most of the RBPs were treated as an outpatient (83%, N=129), hospitalisation was infrequent (17%, N=26, including one overnight stay). Seven outpatients and one hospitalised patient were treated at another ED within the same city even though the GPC recorded a referral to the adjacent ED. Of the RBPs at least 62% underwent lab tests, 14% had conventional X-ray, 38% had other imaging (such as CT scan), 9% had ECG testing, and 14% underwent hospital procedures. Half of the RBPs were administered at least one drug at the ED (31 received painkillers, 7 antibiotics, 3 enema, 13 other drugs, and 26 more than one drug).

**Table 4:**
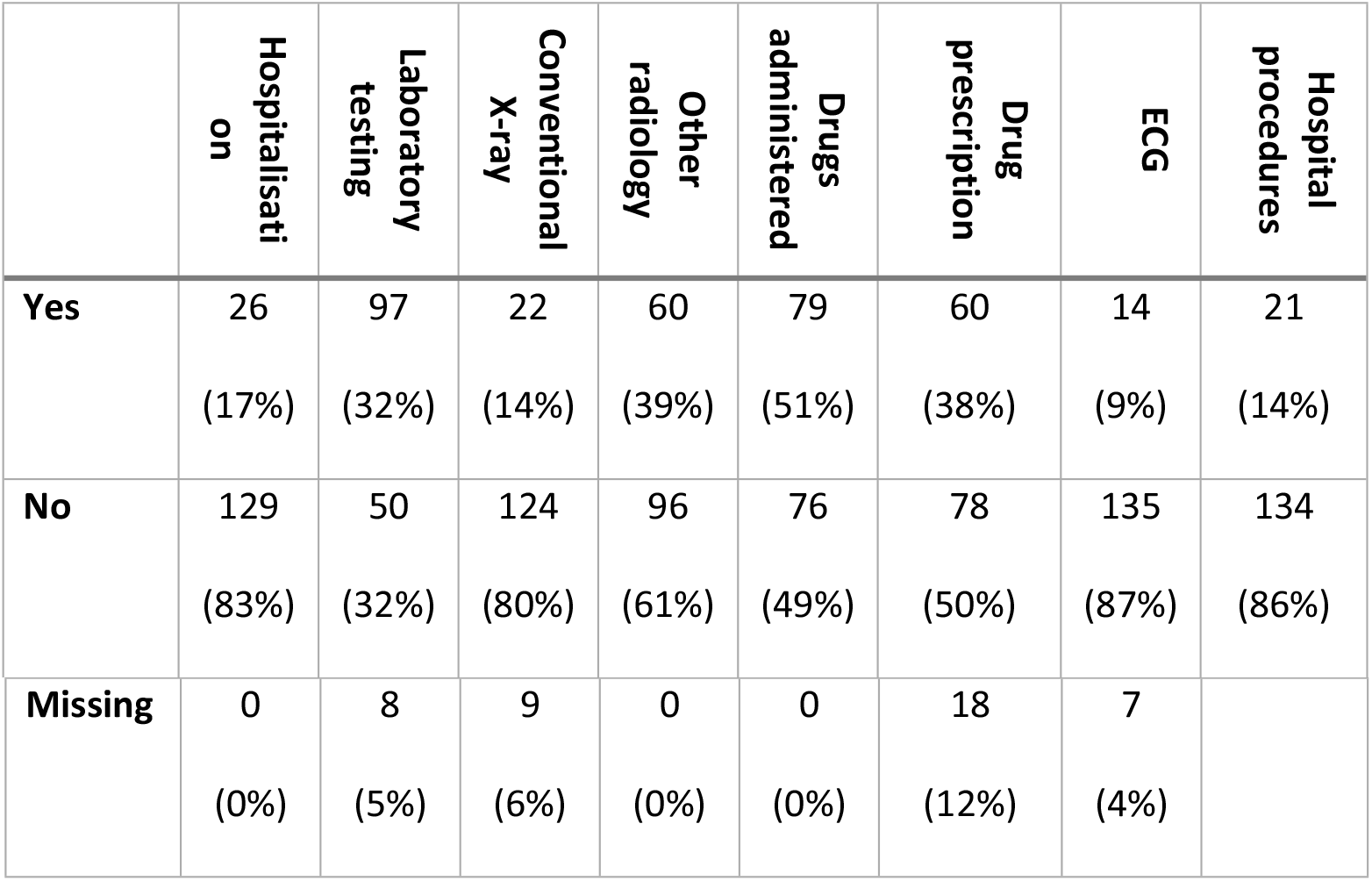
Delivered hospital care in the case review study (155 patients).

Forty-three percent of the RBPs received a drug prescription (26 painkillers, 15 antibiotics, 8 drugs for the gastro-intestinal system, 6 other drugs and 5more than one drug). All prescribed drugs could have been prescribed by a GP as well.

The GP referred 29 patients back to the ED to exclude appendicitis. Twenty-one did not undergo surgery but were diagnosed with other conditions such as viral gastro-enteritis and constipation. Seven cases of appendicitis were confirmed after surgery, in one patient a teratoma was found during surgery.

In four RBPs a major triage issue was found: one patient had instable angina (not treated due to treatment restrictions recorded as DNR2 (Do Not Resuscitate), one patient was in active labour (she was unaware of her pregnancy), one patient had an occlusion of the common iliac artery requiring urgent treatment and one patient had non-ST-elevation myocardial infarction. For 18 RBPs, a minor triage issue was detected. Their diagnoses can be found in S3 Table.

## Discussion

This 6-year follow up study of a RCT conducted in 2019 with the aim to divert low acuity patients from the ED to an OOH primary care setting, revealed an average diversion of 6% of the ED population. This proportion decreased during the COVID-19 pandemic to 3% but afterwards increased up to 11%.

In terms of urgency categories, the referred population was comparable to the general GPC population, with 1% less contacts categorised as “urgent” and none as “highly urgent” by the attending GP, and with half of patients considered as not in need of OOH care.

Three percent of diverted patients were referred back to the ED after evaluation by the GP; this percentage is less than in the average GPC population (5%). A large majority of back-referred patients received an outpatient treatment at the ED and were not admitted to the hospital (83%). Based on a detailed study of the group of back-referred patients (a considerably larger number than during the original study due to the length of the follow-up period), we have more convincing evidence than we had during the RCT that the studied healthcare intervention is safe. Additionally, only five major triage issues were found: during the TRIAGE trial, we reported one major issue (a fatal ruptured aortic aneurysm), four were reported in the current study.

The most frequent reason for referring patients back (N=29) were digestive complaints especially with the request to exclude appendicitis (which was diagnosed in 7/29 or 24%, one patient had a teratoma, the remaining 21 did not undergo surgery). This positive predictive value of 24% for appendicitis was to the results of a Dutch study concerning children in primary care.[8] Because all patients diverted to the GPC were triaged in the blue or green MTS category, they would have had a target waiting time at the ED of less than two hours. Their triage to the GPC led to a time loss of 13 minutes on average (time of registration to time of consultation with the GPC). After back-referral to the ED, they were triaged again with an urgency of at least yellow (maximum waiting time one hour) under the eMTS. This means no crucial time was lost for these patients. This is confirmed by our case review study: none of these appendicitis cases were judged as triage issues. However, patients, ED staff and surgeons are not in favour of transferring patients in and out so when it comes to local support for the triage system, the eMTS flowcharts for abdominal pain should be revised.

These positive follow-up results of our original RCT are not self-evident; RCTs are designed to maximally control the environment with strict inclusion/ exclusion criteria, standardised protocols, and intensive monitoring, and even for pragmatic RCTs, these circumstances are very different from daily routine. Assuming trials with a positive outcome seamlessly transition into implementation within participating healthcare systems overlooks the many factors that must be addressed after the trial concludes.[9]

Frameworks from implementation science provide tools to identify factors that will influence whether an intervention tested in a trial will be sustained in routine practice. Early application of such tools, from the design of the study, helps predict and explain the success of an implementation in real-world settings.[10] Engaging stakeholders in an early phase correlates with higher sustainment, [9, 11] and including an explicit post-trial implementation plan in the protocol is strongly recommended by the World Health Organisation.[11]

Empirical data of recent studies reveal key factors in post-trial sustainment of health interventions: adoption is more likely when an intervention aligns with reimbursement; furthermore, a mismatch between study and routine resources is a barrier to implementation,[9, 12] and other factors than the primary outcomes measured in studies, such as staff satisfaction, may influence the continuation of an intervention.[9]

At the time we designed our RCT, we did not have an explicit long-term implementation plan, which is not in accordance with current standards. However, our pragmatic RCT was designed involving all stakeholders, embedded in routine clinical practice and integrated into routine software to make data collection over a period of 10 months feasible. The studied triage system was not designed for research purposes but originated from a grass root initiative to improve local collaboration. All these factors have likely contributed to the positive long-term results of the trial years after its conclusion. On the other hand, during the trial, a substantial budget was available for training healthcare personnel, and for daily support and monitoring. For example, funding was present to support an extra ED nurse, to compensate for the extra work all ED nurses had while performing and documenting triage. These resources were stopped when the study period ended. Despite the loss of these additional resources, the effects of the intervention were sustained.

Staff satisfaction was a secondary outcome in the TRAGE trial; overall, medical staff stakeholders (ED physicians, GPs and ED nurses) experienced the intervention as positive. ED nurses carried the largest burden but despite the increased complexity, duration, and workload associated with the extended triage protocol and GPC referral for triage nurses, they stated that these measures contributed to a lower (perceived) overall workload.[7] The positive perception of the triage nurses may also have contributed to the durable effects of the trial.

With respect to reimbursement, we found no income loss for the ED in the TRIAGE trial; the revenues of the GPC increased by 13%; the cost for patients was 8% lower but overall public cost was 3% higher.[13] Since the driver of triage in our model is the ED, the absence of revenue loss may be an important facilitator. We were unable to study the long-term financial impact on the ED, because the primary outcome increased after the trial, an income loss is still possible. The lack of overall cost reduction is not necessarily problematic; although cost of out-of-hours primary care is important, numerous other goals such as limiting productivity loss and improving access to and continuity of primary care, are important from both health system and patient perspectives.[14]

Previous evidence suggests that physical triage interventions can safely divert a proportion of ED usage to primary care and can thereby optimise ED performance, [15] however, only short-time data are available. Particularly safety outcomes, cost shifts, and patient trust over time are under-reported. The PERSEE study in the Walloon part of Belgium demonstrated the feasibility of physical triage, with strong algorithmic support as an enabling factor, and with the observation that sustainability depends on algorithm refinement and ensuring safety/confidence in the triage process.[15] One study in Finland reported that over a 13-year period, a combination of interventions (ABCDE triage, public education, reverse triage) could divert young people from ED use and increase the use of primary care during office hours. This suggests that physical triage and referral, combined with systemic changes and public engagement can produce durable effects.[16]

Strengths of our study include the long follow up period of 6 years, which is crucial to measure sustainability and safety, and the use of routine, real-life data. We are one of the first studies measuring long time follow up data. Limitations of our study are the fact that this long follow up was not pre-designed, which results in the fact that we lack information on some of the variables measured in the original RCT. There was no longer a control group available. Additionally the data on care outside of the studied sites is missing so it remains unknown what happened to patients assigned to the GPC but not seen at the GPC. Finally, the case review study was carried out by only one researcher.

## Conclusion

Six years after conclusion of an RCT studying a nurse-led physical triage tool, the tool is still in use, despite the termination of resources associated with the RCT. The efficiency of the tool augmented over the years, and the long follow up period increases the body of evidence regarding the safety of the triage process. To improve the post-trial sustainability of a studied health intervention, it is highly recommended to have an explicit post-trial implementation plan at the start of the study. Abdominal pain and the exclusion of appendicitis during triage are remaining challenges for the effective diversion of ED patients to primary care.

## Data Availability

Due to the lack of individual consent, the authors are not allowed to share the studied dataset. The authors are, however, able to deliver a selection of variables and the outputs of their statistical software upon reasonable request. Such a request should be directed towards the authors.

## Acknowledgments

The authors would like to thank the staff of the study ED an of the GPC for their help, especially Paul Adriaenssens (manager of the GPC) and Jonas Aerts (head nurse of the ED).

Generative AI (Microsoft Copilot 365) was used to improve grammar and readability of certain paragraphs.

## Supporting information captions

**S1 Supplementary Table 1: Reason for encounter in the primary outcome group**

**S2 Supplementary Table 2: Diagnoses in the primary outcome group**

**S3 Supplementary table 3: details concerning 18 patients with a minor triage issue**

